# Knot Guilty? An Examination of Testicular Torsion Litigation Trends from 2014 to 2022

**DOI:** 10.1101/2024.06.30.24309735

**Authors:** Rahma Menshawey, Esraa Menshawey

**Author notes:** Corresponding Author, Email address, Faculty of Medicine, Kasr al Ainy, Cairo University, Geziret Elroda, Manial, Cairo, 11562, Egypt.

## Abstract

**Background:** Testicular torsion is a true urological emergency that occurs when the testes twists around the spermatic cord, cutting off its blood supply. The failure to diagnose testicular torsion is a common medicolegal pitfall due to the uncertainty and urgency of the diagnosis and the potential for testicular loss. In this study, we examined the current medicolegal cases that involved testicular torsion using the LexisNexis database for all cases from 2014 to 2022, using the search terms “testicular torsion” and “medical malpractice”. Our final examination included a total of 20 cases.

**Results:** Trends reveal that Emergency doctors and urologists are the most commonly named defendants. Adults and incarcerated persons are common plaintiffs. The average time from presentation to diagnosis of testicular torsion was 8 ±13 days. The right testicle was the most commonly implicated, and a misdiagnosis was a commonly cited. The average time from testicular torsion diagnosis to filing a case was 4.35 years.

**Conclusions:** Trends reaffirm that testicular torsion remains a high risk of litigation diagnosis, and continued training and education may be needed to remedy the medicolegal pitfalls for this emergency condition.

## Background

“… It is not necessary to practice defensive medicine, but sound medical practice is defensible.(Matteson et al., 2001)”

Testicular torsion is a true urological emergency, which occurs when the testes twists around the spermatic cord, cutting off its blood supply. It requires early surgical intervention in order to potentially salvage the testes as the viability of the testes is directly related to the duration of torsion (Laher et al., 2020).

Testicular torsion is an acute area of litigation due to many factors including uncertainty of the diagnosis, urgency of the diagnosis, and the potential for testicular loss which results in permanent damage and further complications (Matteson et al., 2001). The purpose of this study was to identify and characterize the modern cases of testicular torsion across the time period of 2014 to 2022, and to determine the current trends in the medicolegal landscape involving testicular torsion, based on cases filed in the United States.

## Methods

We searched the LexisNexis Uni Database for “testicular torsion” and “medical malpractice” cases from January 1, 2014, to December 31, 2022. The database contains court cases and legal news, commonly used in medicolegal research. After screening the search results, we identified 33 cases. We excluded cases where defendants were not healthcare entities or practitioners. Data collected from legal documents included the number and specialties of defendants, plaintiff’s age (adult or child), affected testicle, time from complaint to torsion diagnosis, time from torsion to case filing, states where cases were filed, use of ultrasound, and relevant medical and legal issues, including expert witness opinions. (see figure 1)

**Figure 1:**
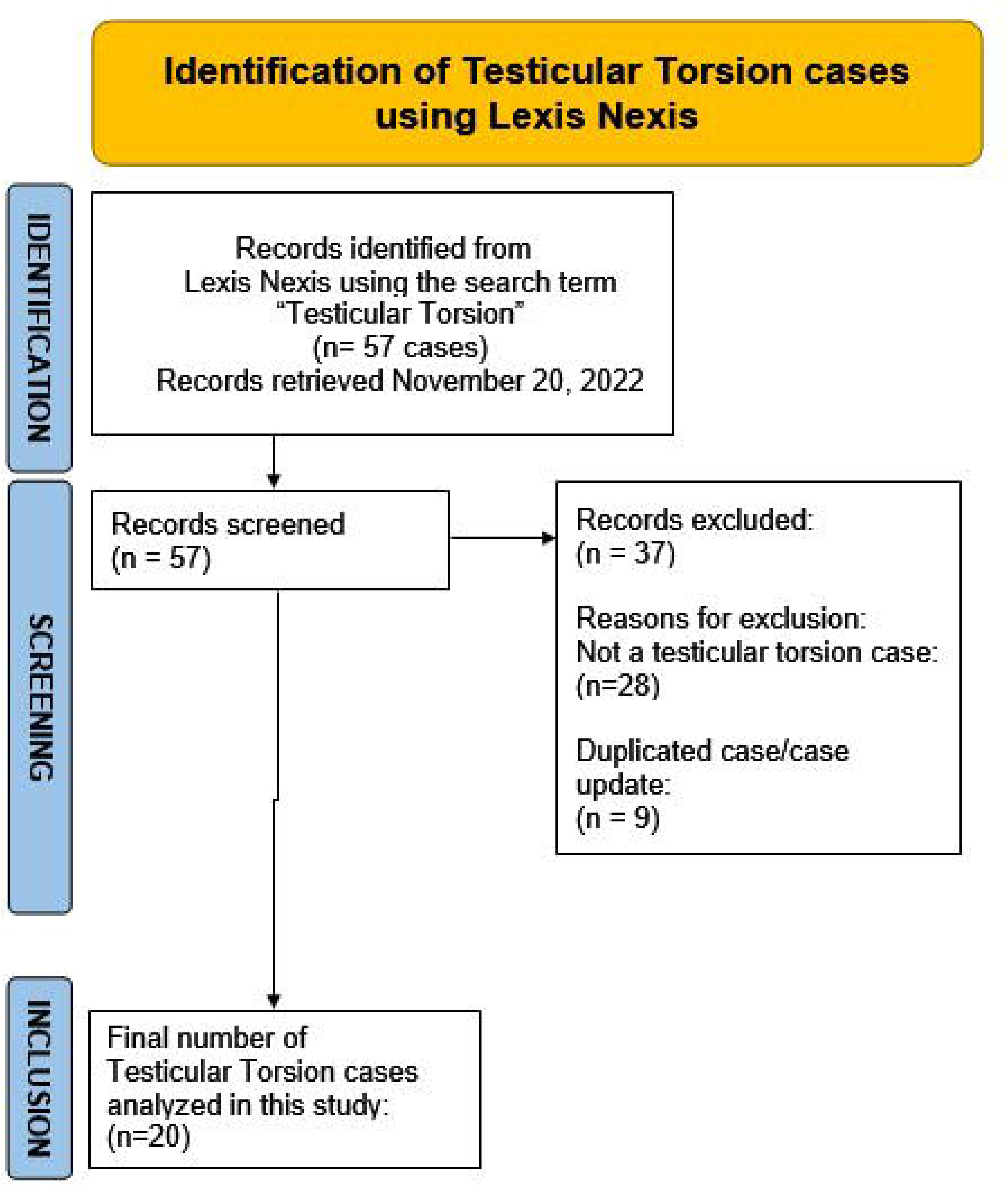
Flow chart illustrating the method for identifying relevant cases in the Lexis Nexis database. After screening for relevant cases and excluding duplicates and non-testicular torsion cases, 20 cases remained for examination.

## Results

A total of 57 cases were identified on the Lexis Nexis database. The final analysis was performed on a total of 20 cases. The cases were filed across a variety of states (see figure 2). The majority of testicular torsion cases were filed in the state of New York and Pennsylvania, 25% (n=5) and 15% (n=3) respectively. 45% of the cases examined involved a plaintiff that was a prisoner (n=9). 70% of the cases involved a plaintiff that was an adult (n=14), while the remaining were children or infants. 55% of cases involved torsion of the right testicle (n = 11), while 30% involved the left testicle (n=6), and in 3 cases the involved testicle was not mentioned in the court documents.

**Figure 2:**
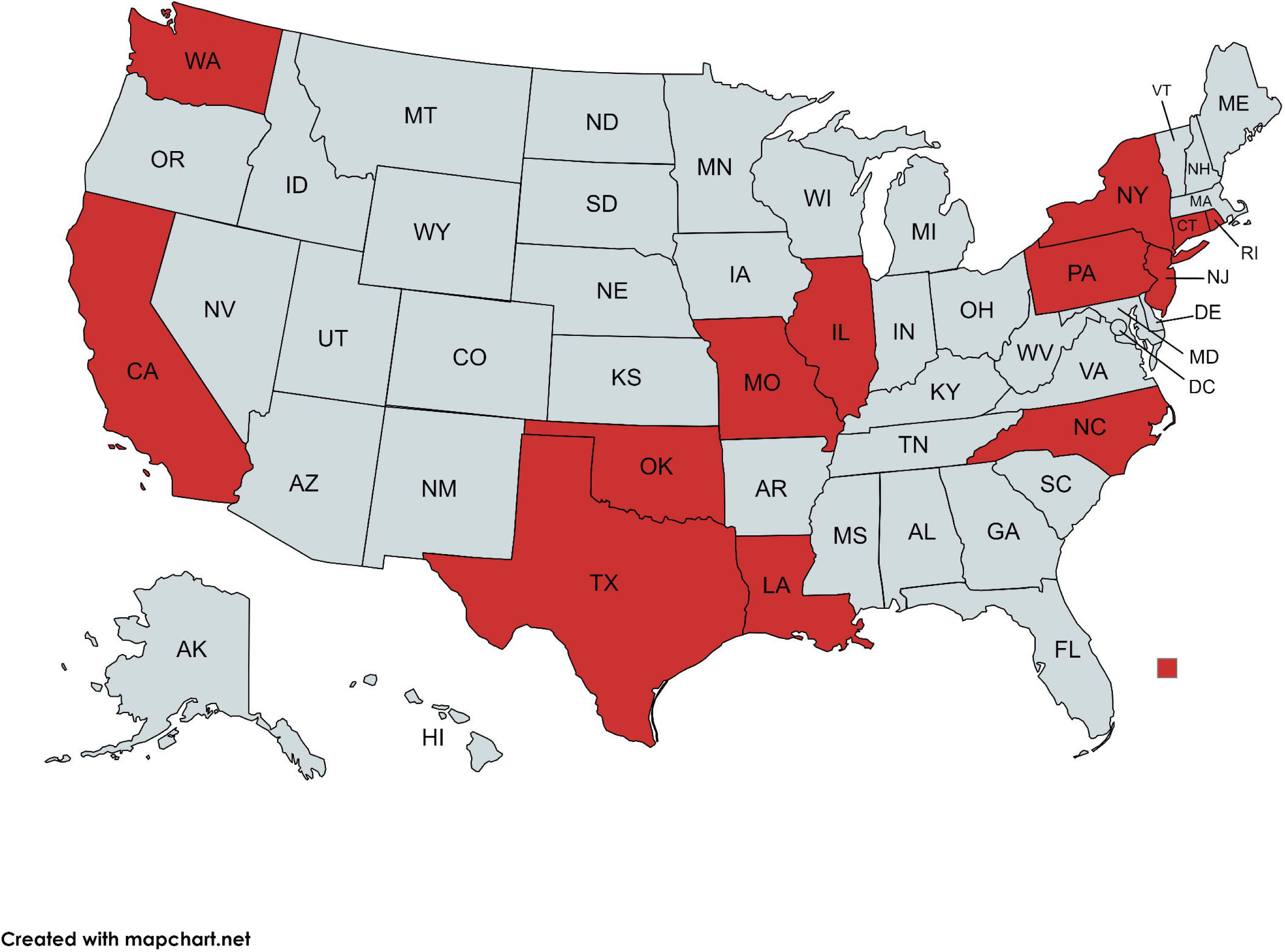
States where the cases were filed. New York had the highest number of filed cases.

Several medical practitioners and medically related defendants were implicated in the filed cases, including physicians, nurses, nurse practitioners, physician assistants, technicians, hospitals, and hospital networks. Among the 20 cases, a total of 70 medically relevant defendants were listed. MDs accounted for 47.12% (n=33), while Nurses/NPs accounted for 28.57% (n=20) (Figure 3). Among MD defendants with identifiable specialties, 30% were Emergency physicians (n=10), and 12% were urologists (Figure 4) (see Table 1).

**Figure 3:**
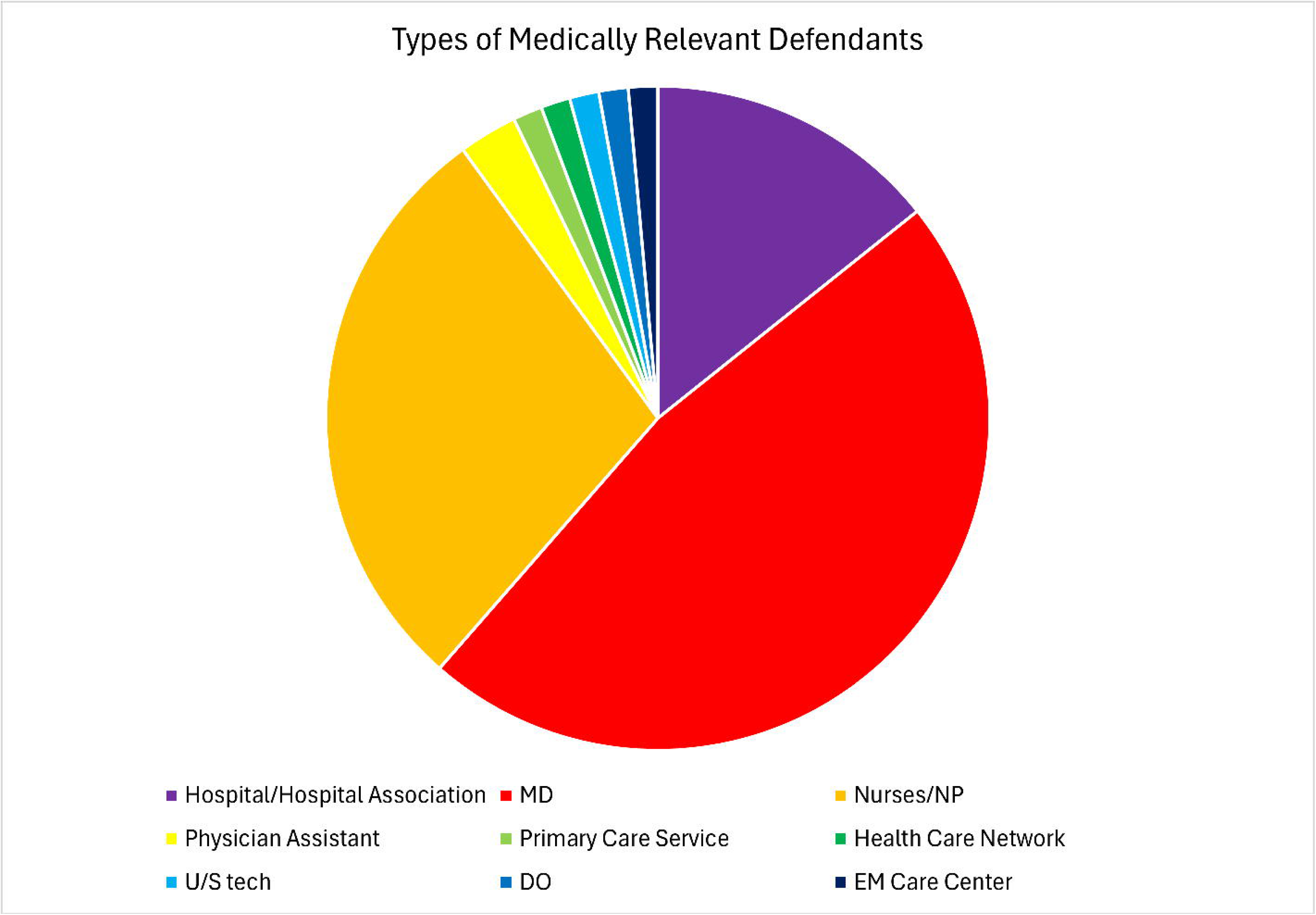
Types of defendants. MDs and Nurses were primary defendants.

**Figure 4:**
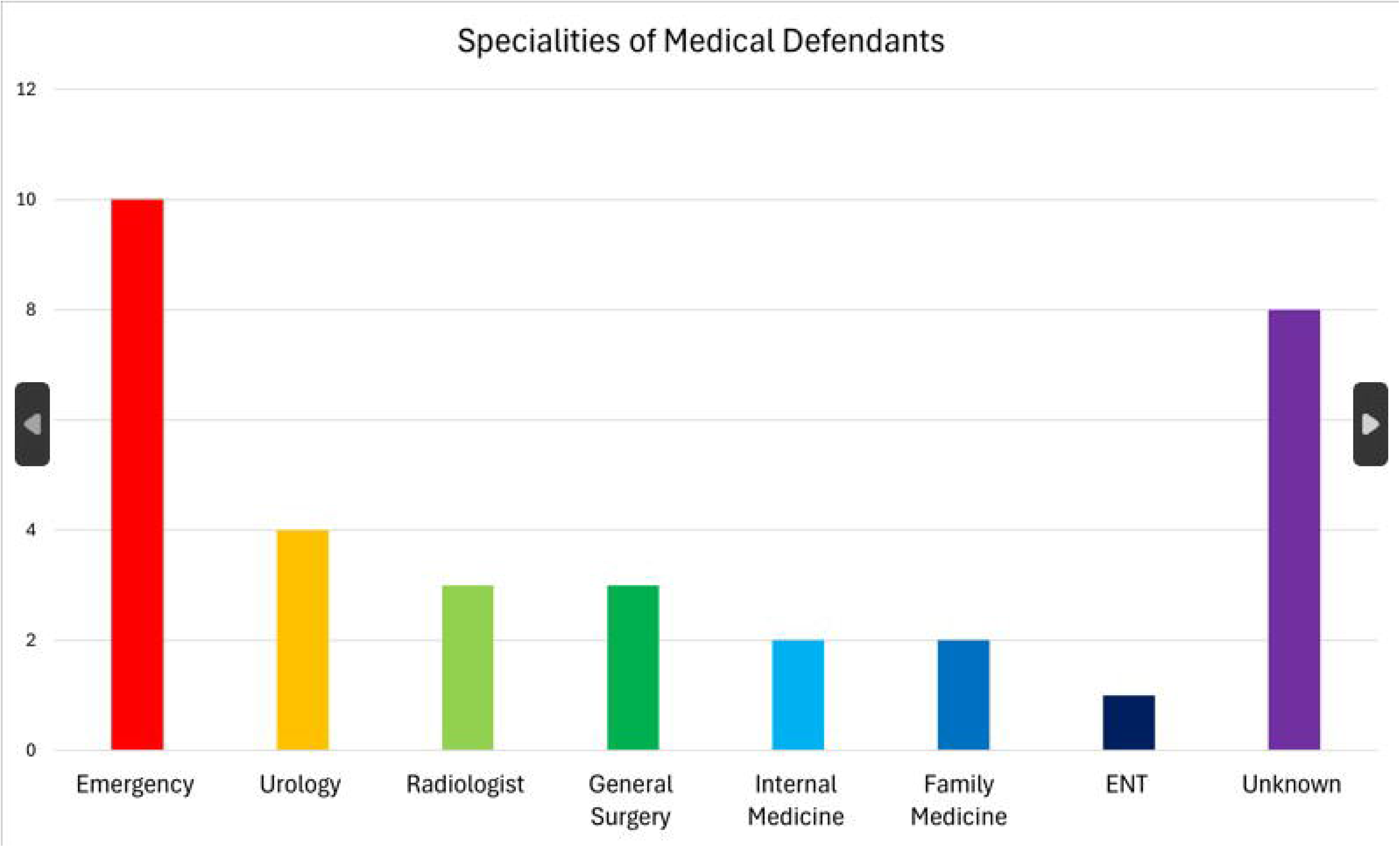
MD defendant specialists. Consistent with previous trends, emergency physicians and urologists were commonly named as defendants.

**Table 1:**
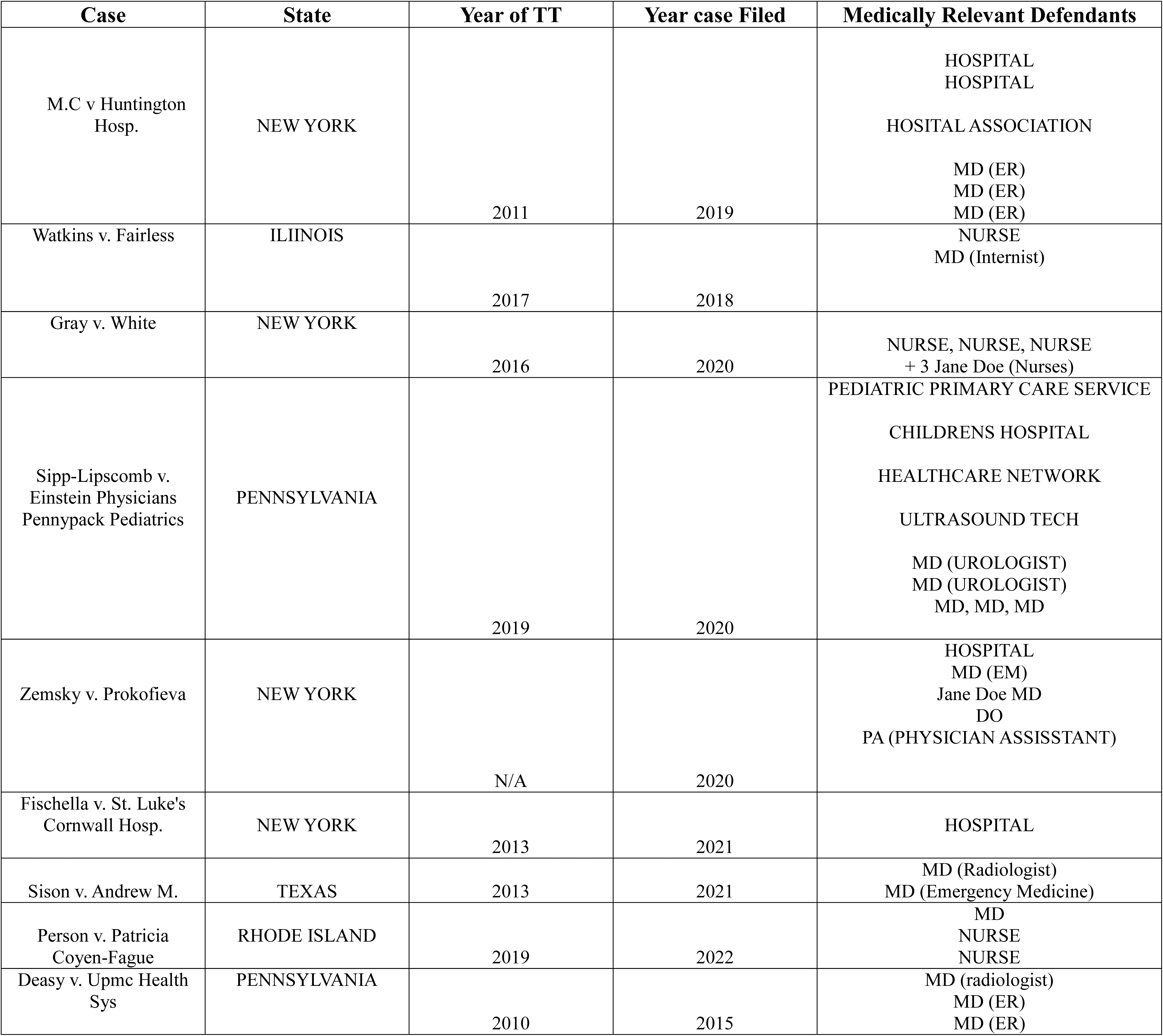
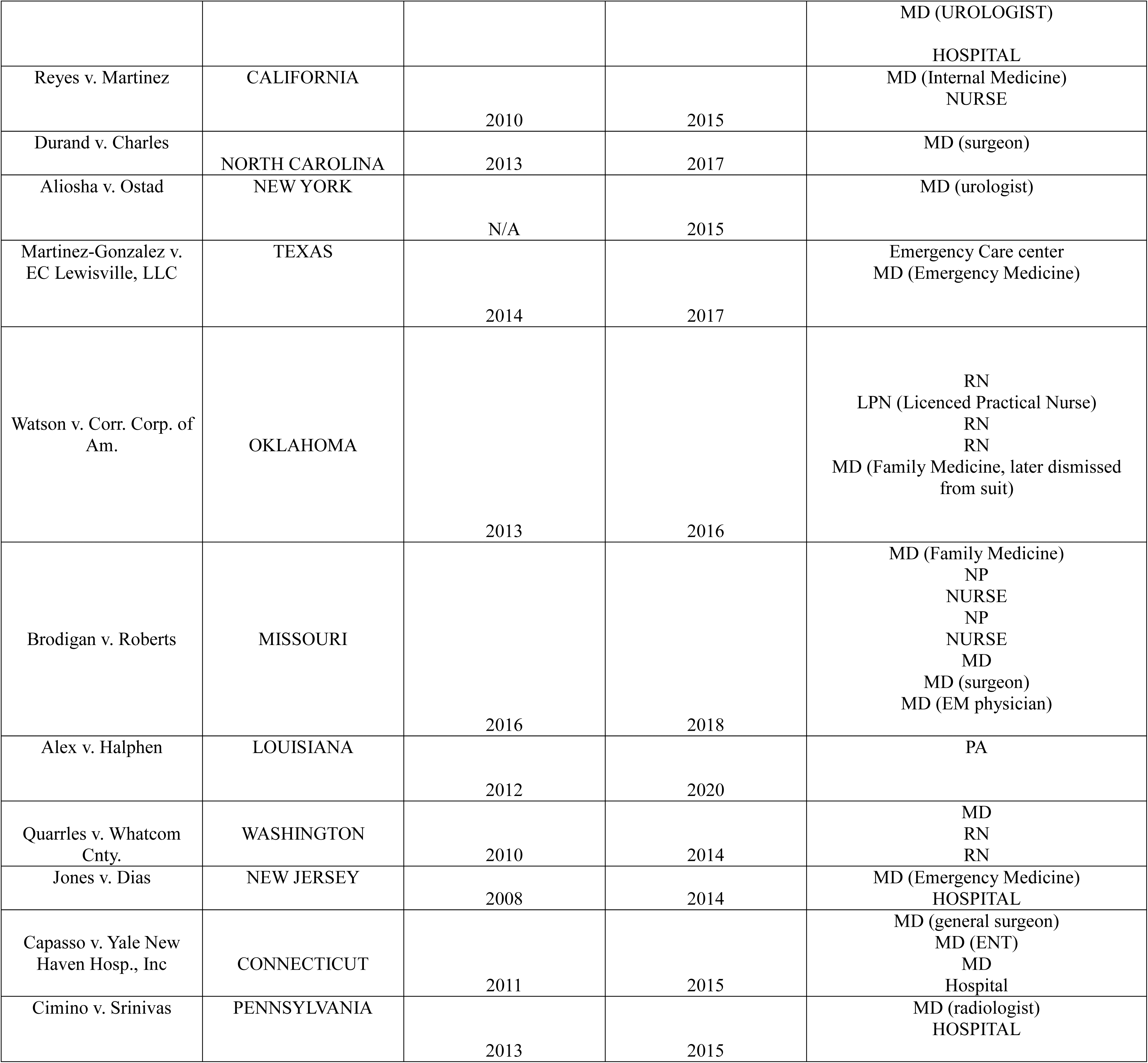
Testicular Torsion Litigation Cases Details and Medically Relevant Defendants.

Ultrasound usage was mentioned in 55% of cases (n=11), was not used in 35% of cases, and was not mentioned in 2 cases. Only 20% of cases explicitly mentioned a urological consult. Among cases which specified dates, the average time from symptom onset to testicular torsion diagnosis was 8 ±13 days, with the longest being 48 days and the shortest being 1 day (see table 2).

**Table 2:**
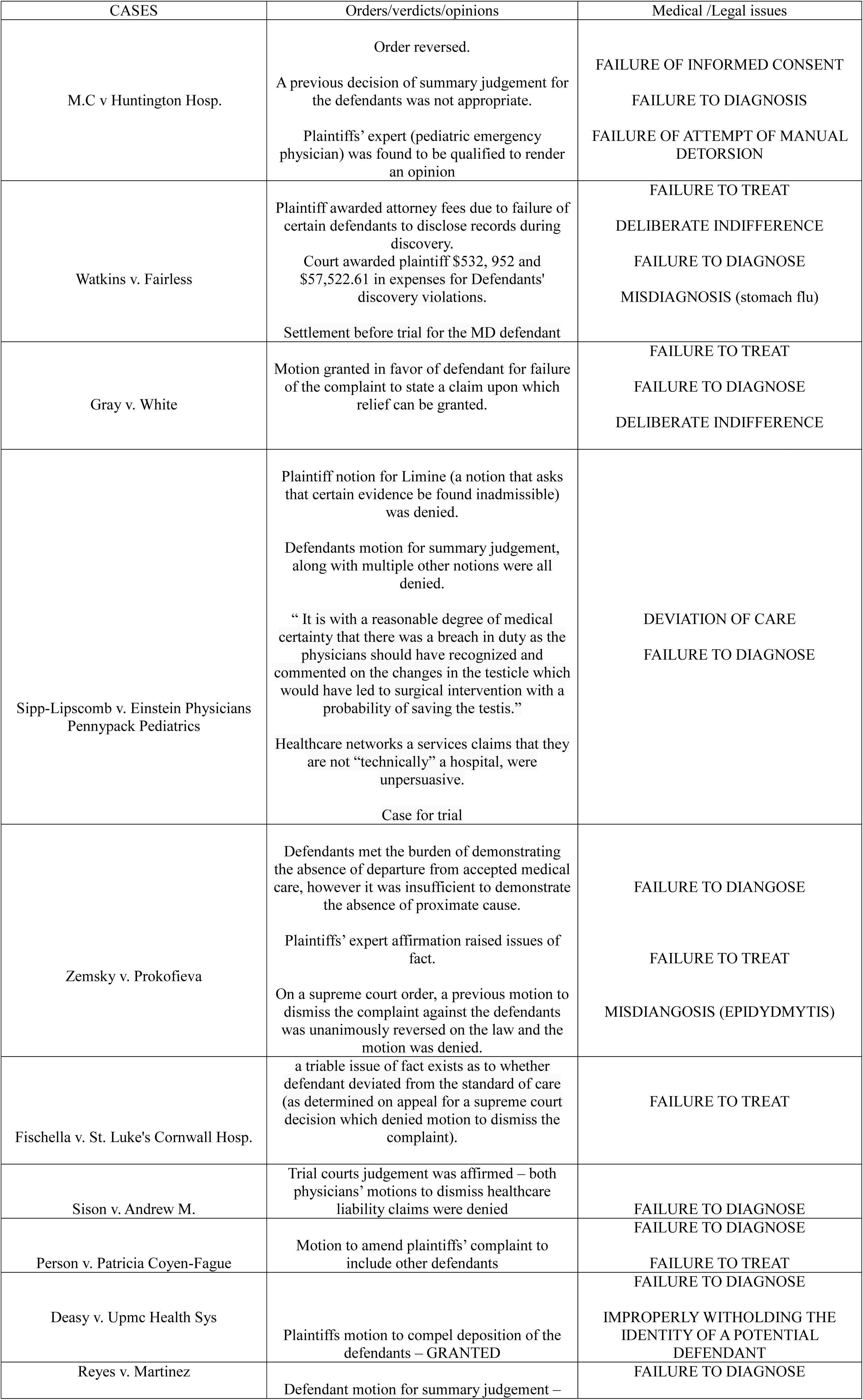
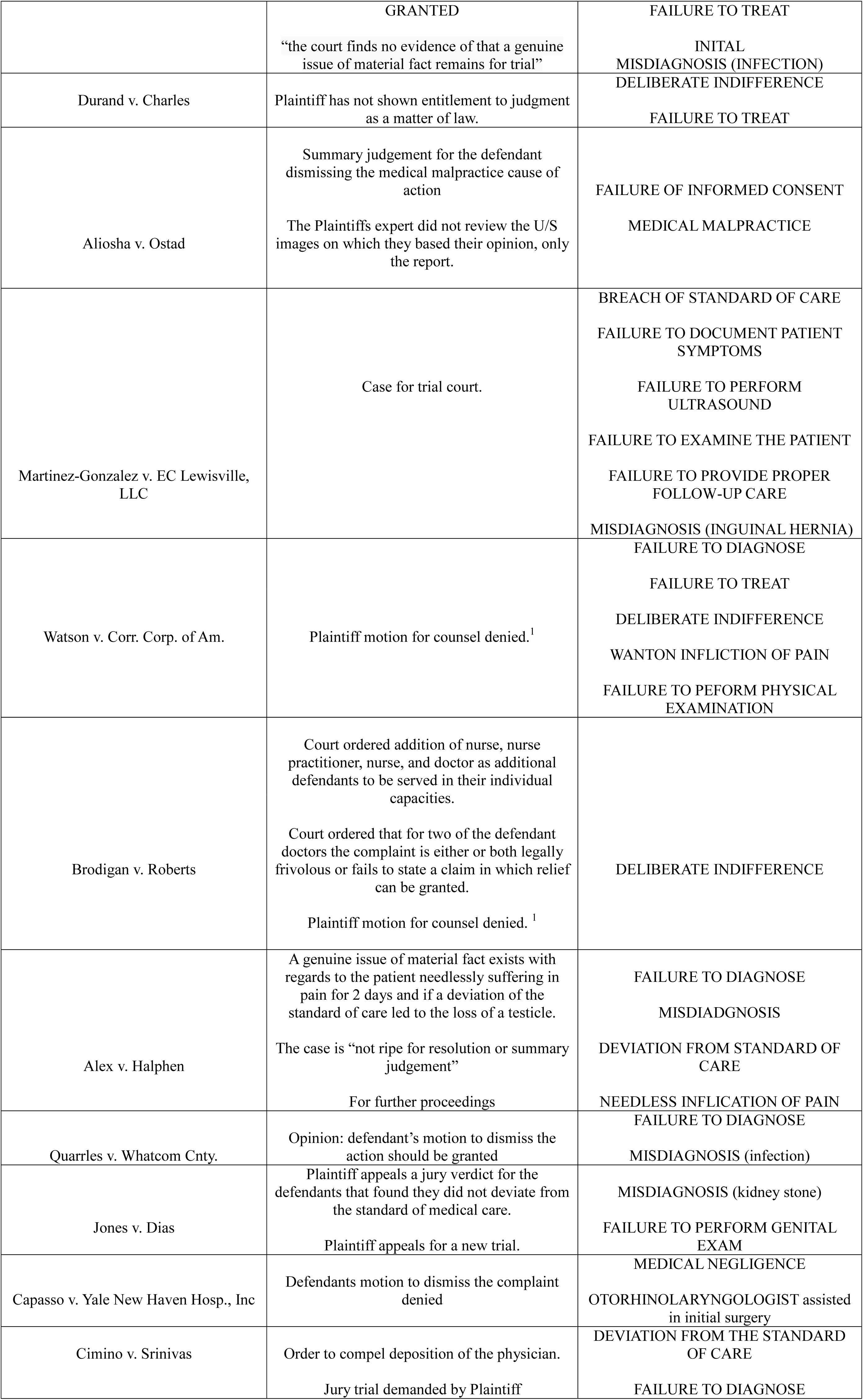
Medical and Legal Outcomes of Cases.

For cases indicating both the year of torsion occurrence and case filing, the average time between these events was 4.35 years. The longest time from torsion to case filing was 8 years, and the shortest was 1 year (Figure 5).

**Figure 5:**
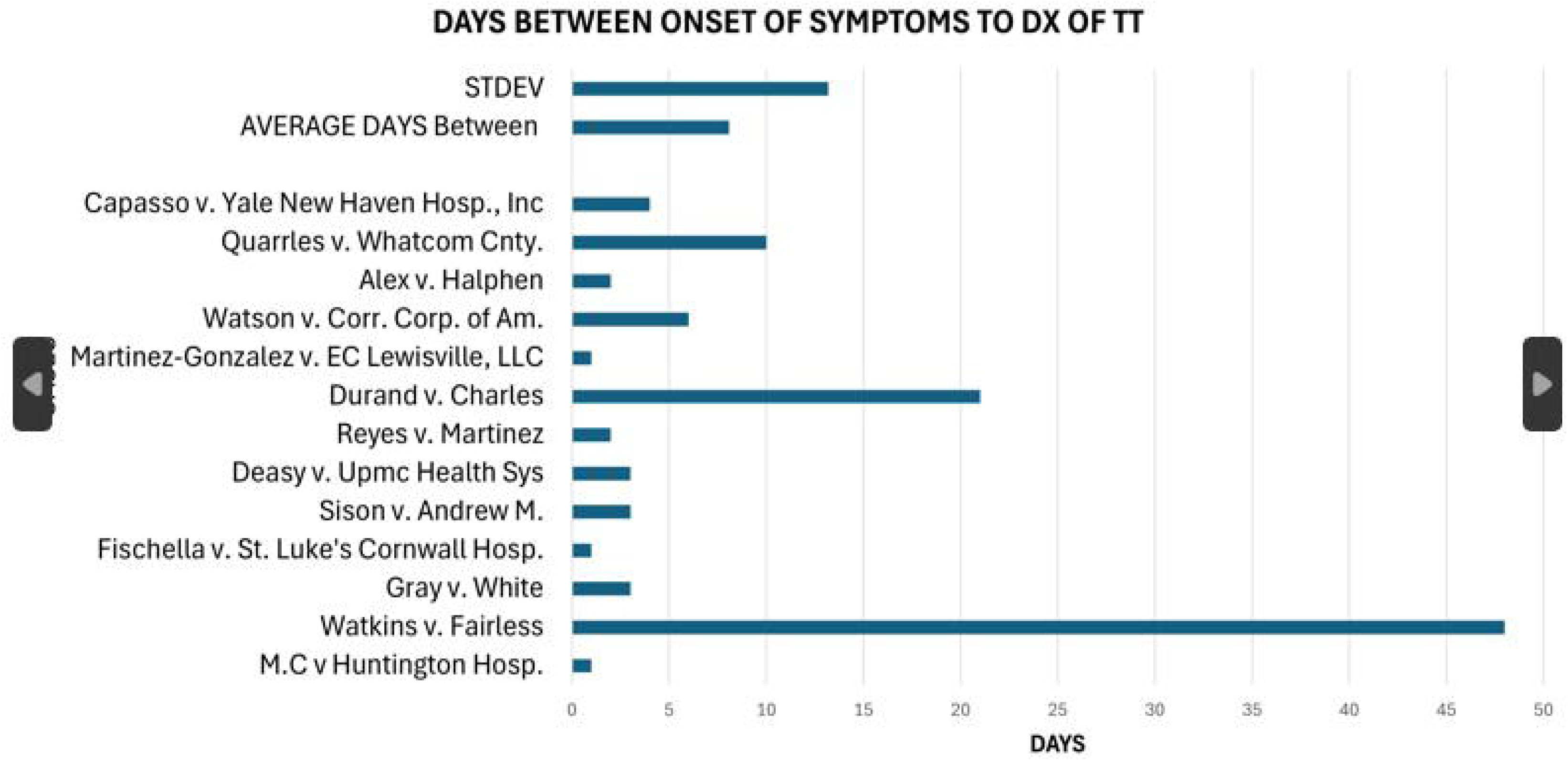
A cluster graph representing the time from symptom onset to the diagnosis of testicular torsion in the cases.

Multiple medical and legal issues were identified, including deliberate indifference, failure of informed consent, failure of diagnosis, misdiagnosis, failure to perform ultrasound and examination, inadequate follow-up care, deviation from standard care, infliction of pain, surgical assistance from unrelated specialists, and deliberate withholding of information or identity of defendants (see table 3). A total of 52 explicitly mentioned issues were identified across the cases, averaging 2.6 issues per case. Misdiagnosis was explicitly mentioned in 35% of cases (n=7), with infection/epididymitis as the most common misdiagnosis, along with stomach flu, kidney stone, and inguinal hernia. We provide a narrative summary of the medically relevant context of each case in a supplementary file.

**Table 3:**
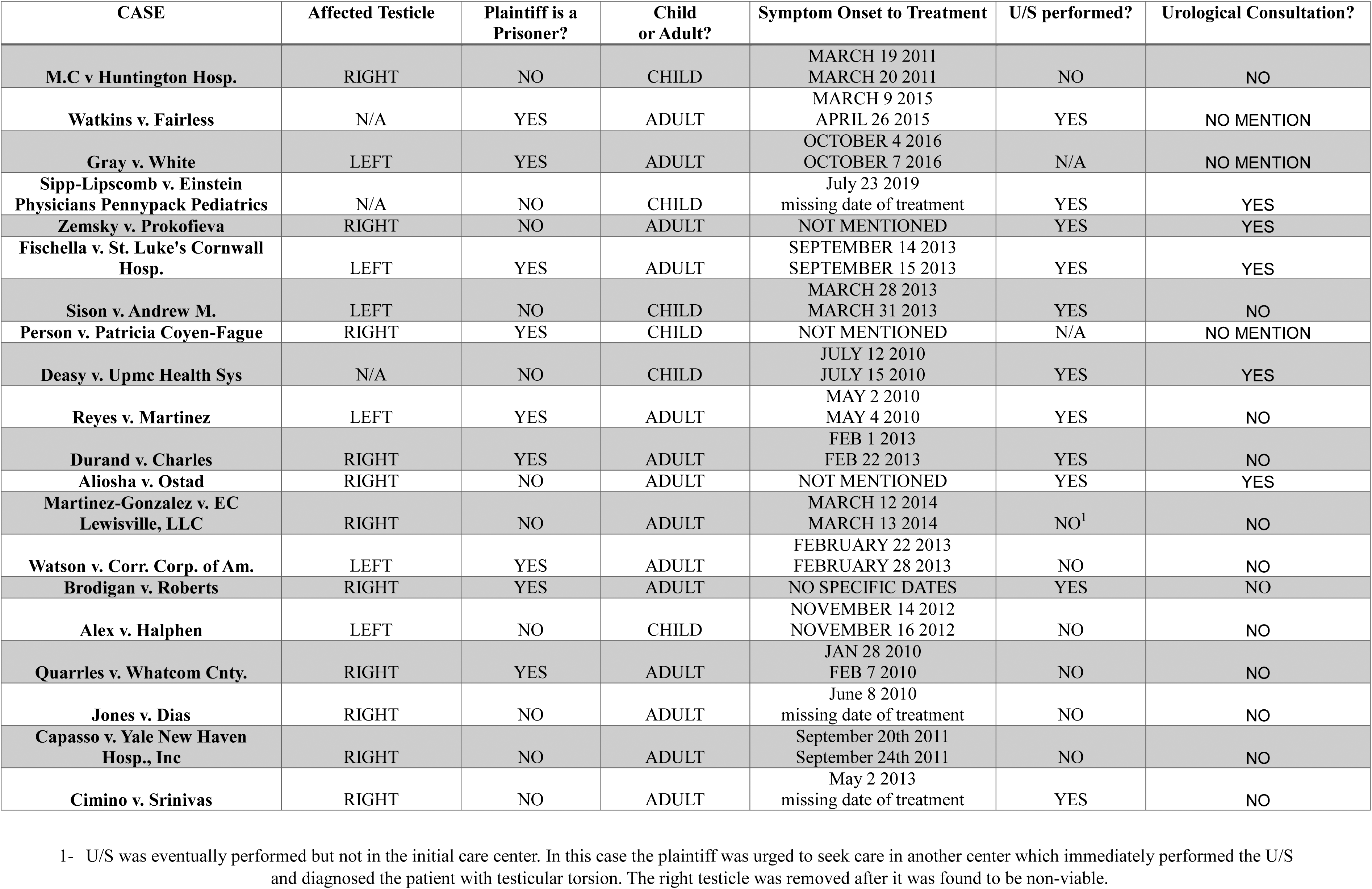
Medical and Legal Outcomes of Cases.

## Discussion

### Results and Trends

We examined testicular torsion malpractice litigation from 2014 to 2022. Interesting trends emerged from our analysis. Most cases involved adults, with the right testicle being the most commonly affected. Urological consultations were infrequent in the examined cases. The majority of cases had multiple medically relevant defendants, with physicians and nurses being the most common. Emergency physicians and urologists were the leading specialists named as defendants in testicular torsion litigation.

These trends can be explained by the fact that testicular torsion primarily affects pediatric patients. Consequently, in adults, it may not be prioritized in the differential diagnosis, leading to failures or misdiagnoses with conditions like epididymitis or kidney stones. Additionally, cases involving the left testicle are more common, resulting in potential misdiagnoses of right-sided cases and explaining the higher number of right testicle-related cases.

### Testicular Torsion and Medical Malpractice

*“…*physicians involved in litigation for arguably mismanaged cases of testicular torsion may be at risk for increased financial liability relative to other medical malpractice cases, and the window of concern may be prolonged.” [3]

There are 4 key elements to claim medical practice:

1. The physician has a duty
2. The physician has breached that duty
3. A harm has resulted.
4. And that harm is directly due to the aforementioned breach.

Points 1 and 3 can be easy to prove, while points 2 to 4 can be more challenging [4]. The plaintiff must prove that there was a breach in a standard of care, and that this breach resulted in an approximate harm.

Testicular torsion is an area ripe for litigation due to delays in treatment and diagnosis, and the impact on reproductive function. Indemnity payments are common, and legal outcomes may not be affected by late presenting torsion. Error in diagnosis and lack of or delayed referral are commonly cited causes for liability (Matteson et al., 2001). Testicular salvage studies seem to reaffirm the “6 hour rule”, and so surgical exploration within this time frame maybe considered “legally safe”[6]. Hospitals and physicians may be held liable for the delays under their control, and there are other hurdles to consider such as the patient not immediately presenting on the onset of pain, or seeking care with a general practitioner first, and the time needed for investigations, consults, and availability and preparation of an operating room [6]. Mismanagement or initial misdiagnosis maybe the trigger for litigation as in the cases we examined the named defendant was almost always the first encounter the plaintiff had with a healthcare provider (be that a nurse, physician assistant or physician), and did not often include the surgeon or urologist who actually diagnosed and removed the dead testes. This could explain the rise in naming emergency physicians in lawsuits more commonly over urologists. Urologist or surgeons were named as defendants if the mismanagement or misdiagnosis involved them, like failing to recognise no flow on a scrotal ultrasound. Expert statements by certified urologists are a pivotal part of the cases for both plaintiffs and defendants and often clashed at whether a deviation of care occurred, whether an ultrasound showed testicular torsion, and the decision to proceed with US and further consultation instead of directly proceeding with surgery based on the patients’ initial findings and symptoms. The established idea is that testicular torsion is a true surgical emergency, and that surgical exploration is the only true way to diagnose it is often cited by experts for the plaintiff. This also explains another trend in the litigation literature where having obtained an ultrasound had no bearing on the judicial outcomes. Another area of clash among the expert opinions involved the topic of misdiagnosis.

Literature review reveals interesting findings regarding testicular torsion litigation.

Matteson et al examined files from a large medical malpractice insurance company across 1979 to 1997. Indemnity payments were made in 67% of cases with a median payout of $45 000. Urologists were the most names specialists, and a misdiagnosis (epididymitis) was commonly cited. Late presentation did not affect medicolegal outcomes (Matteson et al., 2001).

Colaco et al examined cases from 1990 to 2013 and found that 51% of trial verdicts were in favor of the defendant across those decades. Emergency medicine doctors accounts for 48% of total defendants while urologists made up 23% of the defendant pool [3]. Among urologists, cases were unsuccessfully defended if the defendant was a resident. No significant differences were found in between ultrasound users and nonusers in the rate of a successful defence. Most common misdiagnosis were epididymitis, gastritis, orchitis, UTI, and kidney stones[3].

Another study examined state appellant cases involving TT from 1985 to 2015. State appeals were in favor of the provider in 50% of cases. Emergency providers were the most commonly sued. Atypical presenting TT was common. Providers who used an US were not more likely to win a state appeal [7].

Clennon et al examined the literature involving testicular torsion and ovarian torsion, and they found that malpractice cases were 38x more likely to arise from testicular torsion cases [8].

One Spanish study examined medical liability claims from 2000 to 2018. Complaints were reported on first consultation. U/S was only performed in 7.5% of cases [9]. Another study examined cases across Spain and France. The most common misdiagnosis was Orchiepididymitis and gastritis or abdominal pain. Urologists were the most frequently implicated specialists in both countries. It was recommended that proper filling of emergency reports and medical records is essential for defending possible malpractice claims [10].

One study examined 2 additional cases. In one case a patient was discharged from the ED after an unremarkable abdominal and pelvic CT. Genital examination was not performed. The next day the patient returned to the ED and the testicle was removed. A settlement was reached at $300 000. In the next case a patient arrives to the ED and appendicitis is ruled out by CT (surgeon consult), and TT ruled out incorrectly by a radiologist consult on scrotal ultrasound. The emergency physician was sued, and the case did not name the radiologist. The logic in this case was that the Emergency physician should have been able to make a clinical correlation that the radiologist could not. The jury awarded verdict for the plaintiff and a $500 000 award. The authors stress the following points [11]:

- Presentation circumstances may not always indicate the diagnosis of testicular torsion (TT). For instance, TT can occur during sleep due to spontaneous cremasteric contractions. Relying solely on historical features may be insufficient.
- Despite ultrasound (US) being considered a “gold standard,” it is not infallible. If clinical suspicion of TT remains high, a consultation with a urologist is necessary. Seeking a consult establishes a strong defensible position.

## Conclusions

This study has certain limitations, including potential delays in case availability on databases due to updates within the court systems. Legal cases often lack medically relevant details that may be more valuable to a medical audience. Nevertheless, legal databases have been crucial in providing essential information for analyzing medicolegal concerns. In conclusion, testicular torsion remains a high-risk emergency condition associated with litigation. Trends consistently point to emergency physicians and urologists as the most commonly named defendants. Adults and incarcerated individuals are frequently identified as plaintiffs during the examined period. Misdiagnosis remains a significant medicolegal pitfall.

Litigation research on medical malpractice associated conditions, such as our study, can provide insight into areas that require more support such as targeted interventions, with the hope of prevention, patient safety, and the reduction of harm. While the literature is predominated by litigation in the United States, and so global research in this area could arise to new and insightful context. Litigation research can also lead to a better understanding of litigation trends, and this can help to guide the efforts involving patient safety and quality care.

## List of abbreviations

TT: Testicular Torsion
US: Ultrasound

## Declarations

### Ethics Approval and Consent to Participate

Not applicable

### Consent for Publication

Not applicable

### Availability of Data and Material

All data is available in this manuscript and in additional supplementary files.

## Authors Contributions

RM conceived the idea. RM and EM compiled information, analysed the results, wrote the manuscript and approve its final form. All authors have participated significantly to this work to warrant inclusion as an author based on ICJME guidelines for authorship.

## Data Availability

All data produced in the present study are available upon reasonable request to the authors

## Acknowledgements

Not Applicable

